# An exploratory study on becoming a traditional spiritual healer among Baganda in Central Uganda

**DOI:** 10.1101/2024.04.02.23297238

**Authors:** Yahaya H. K. Sekagya, Charles Muchunguzi, Payyappallimana Unnikrishnan, Edgar M. Mulogo

**Author notes:** We have no known conflict of interest to disclose. Email; ORCID: 0000-0001-9721-7158.

## Abstract

Traditional medicinal knowledge and healing practices of indigenous spiritual healers play important roles in health care, and contribute towards achieving Universal Health Care. Traditional spiritual healers (TSHs) are grouped into three categories. One category of Baganda TSHs, *Balubaale*, engage ancestral spirits during health management. *Balubaale* are socially significant but not legally accepted. Their initiation and training practices have not been documented in Uganda. The study purpose was to understand and establish the training of traditional spiritual healers. Twelve (10M, 2F); practicing TSHs in Central Uganda were purposively selected and recruited between 15^th^ July 2019 and 29^th^ April 2020, and were prospectively interacted with for 24 months. Transcribed data was coded and thematically analyzed using ATLAS ti. 22 computer software and presented based on an inductive approach. Findings show key areas of TSHs training include connecting with ancestral spirits and the spiritual powers of non-materials and materials such as living and non-living things through rituals. Spiritual healers train in diagnosis and health management based on ancestral spirits and they finally pass out in a communal ceremony witnessed by family and community members. We conclude that TSHs undergo training and are supervised and supported by experienced spiritualists, family and the community. We recommend similar studies among other ethnic groups to contextualize the process of becoming a TSH, compare and harmonize findings to facilitate inter-medical systems communication and policy considerations.

## Introduction

Spirituality in health care, and the use of indigenous knowledge, beliefs, methods and practices to maintain health of individuals, families and communities are increasingly securing interest of researchers and health workers globally (1–4). However, the lack of scientific knowledge in the beliefs, methods and practices of spiritual healers (5), challenge the regulation of these health providers with low levels of formal training and different therapeutic practices (6,7). Advocacy for multi-perspectival knowledges is currently supported to allow education and research to respond to specific cultural and social needs of diverse populations (8,9) to contribute towards achieving Universal Health Care (10) in culturally appropriate ways (11–13). United Nations Declaration of the Rights of Indigenous People recognises the contribution of Indigenous knowledge, cultures, and traditional practices to sustainable health, education, and proper management of environment. For example, Article 14 and 15 emphasis the rights of Indigenous People to establish and control their educational systems in their own languages, in a manner appropriate to their cultural methods of teaching and learning. Likewise, Articles 24 and 31 encourage them to maintain, control, protect and develop their traditional medicine, health practices and cultural expressions (8). This is supported by recommendations that trained and categorised healers with recognised competencies should be identified, profiled, registered, monitored and evaluated and get certified by accredited institution that understand, contextualize, appreciate, and recognise them and their practices (14,15). More authors argue that this would assist to harmonise, streamline, regulate and promote informal skills development, qualifications, and accreditation of their practices, therapies, and services (16–19). Nonetheless, the practices of traditional healers do not lend themselves to evaluation, measurement, and precise data, using qualitative and quantitative methods (20), even though local and indigenous ways of knowing have long co-existed with mainstream sciences (12).

In Africa, traditional healers diagnose illness, identify sources of problems and find solutions through communication with ancestors (21–23). Although some countries have legalised traditional medicines, they have left out aspects of spirits and spirits possession (24–26), due to limited literature to enable policy makers appreciate their health roles (27–29). The regulation of registration, training and practices of traditional spiritual healers is assigned to traditional health practitioners’ councils (24–26). The WHO recommended and mandated the member states to establish national databases on traditional medicine practices, practitioners, and their respective specific needs to upgrade their knowledge base and skills (13).

In Uganda, 60% of the population use traditional health care (13), for paediatric illnesses (35), psychotic disorders (36–38), gynaecological and general unspecified “African” diseases (39), where spirituality features in many aspects of everyday life (40–42). A health worker is defined as any person who holds a qualification recognized by Uganda government (18). This definition excludes spiritual healers, as human resource for health (13,17,18). This implies that spiritual healers and their clients are not protected by law, and their integration into community and national health structures [29] is denied. For example, the Traditional and Complementary Medicine Act 2019 does not mention traditional spiritual healers (43). However, their current practice is supported by their cultural and clan belief system, its values and worldviews. They are challenged by lack of legal status, which aspect sustains their experience of unfavourable direct and indirect foreign rule (44). Therefore, training and practice engaging ancestral spirits during health management is vulnerable to widespread charlatanism, misinformation, ambiguity (13,45), and no official levels of utilization of traditional spiritual healers in health care (15). It is, therefore, a national challenge and priority to understand and regulate training of traditional spiritual healers, their practices and premises (43) so as to inform policy (23).

There is a major gap relating to lack of systematic evidential knowledge about training and methods of learning of traditional spiritual healers. This study, therefore, focused on Baganda spiritual healers with a natural call for healing others, and explored their lived experiences and the ritualistic processes they undergo to become acceptable in their communities in Buganda – Central Uganda. This article provides a foundation for further research towards policies that promote utilization of local experiential health knowledge in the national health care system.

## Method

### Study design and setting

The study was designed to qualitatively explore the process of becoming a traditional spiritual healer among Baganda in Central Uganda. We used an ethnographic approach (46), and followed step-by-step research procedure (47–49). Uganda covers 241.555 km^2^ and is divided into four regions namely Eastern, Western, Northern and Central. Uganda currently has 156 districts with a population of 44.2 million people of which 88.6% are in rural areas and subsistence farmers (50). The central region (Buganda), which is the focal area of the study, is comprise of 27 districts, and is dominated by Baganda. Baganda are Bantu speaking and the largest ethnic community in Uganda, headed by a cultural leader called Kabaka. Baganda are divided into social or kinship divisions called *bika* (singular *kika*), which means clans. Baganda clans prohibit intermarriages within the same clan and are patriarchal, with strong emphasis on male authority. Each clan is a family group which traces its origin to a common ancestor and has common totems, mostly animals, held sacred by all its members (51).

### Study population and participants identification

The study population comprised of Baganda traditional spiritual healers (*Balubaale*) who worked and resided in Central Uganda. Purposive sampling was used to identify participants who were available, proficient with rich and deep specialized training knowledge (52) and at least ten (10) years of practical experience of training others to become spiritual healers (*Balubaale).* Similar to a qualitative study by Akol et al, participants were identified by leaders of their associations (53). For this study, additional list of names of participants was done by district and cultural leaders. Letters were addressed to District administrative leaders for names of known and experienced Baganda traditional spiritual healers working and residing in their respective districts. Another letter was addressed to Minister responsible for culture in Buganda Kingdom for permission to access clan leaders for such healers.

A final compiled list comprised of 149 names of TSHs. Thirty names could not be contacted and were eliminated. Sixty-eight did not meet inclusion criteria (See figure 1), and were eliminated. During field work, the inclusion criteria was adjusted to include only spiritual healer mediums for ancestral spirit *Muwanga* because it was established that they are more ethical and disciplined, which considerations were very important for the study. Out of the remaining 51 spiritualists, 33 who had trainees were considered to be more practical, were selected and visited at their individual shrines. We introduced ourselves and the study to each spiritualist. We went through prior informed consent word-by-word, and after addressing their concerns, we requested for their permission to participate in the study by signing or thumb printing the prior informed consent forms. Twenty declined with or without giving reasons. One healer was eliminated because his shrine was unhygienic with almost-full single stance pit latrine, which was considered unfit to host the study team. All the remaining twelve (10Male; 2Female) were recruited in the study between 15^th^ July 2019 and 29^th^ April 2020 and were prospectively interacted with for two years. Earlier studies, one by Akol and another by Canel successfully used small numbers of participants and retained validity in their studies (53,54)

**Figure 1.** Study Profile

We selected Baganda adult spiritualists with shrines, and were spirit mediums including for ancestral spirit *Muwanga*. The study excluded spiritual healers who confessed to be of other tribes and those residing outside the study area (55).

### Data collection

Data was collected through semi-structured interviews using open-ended questions (56–58), and partial integration observation method for subjective understanding of lived reality (59).

The first author revealed his orientation and personal involvement in the research (60). Using participant observation, the research team was involved in daily health care activities and interviewed practitioners as appropriate which lessened interference with their usual practices. The research team was trained in note-taking and kept personal self-reflective journal for self-reflections regarding interview responses, assumptions, confusions and bias (61). Interview and observation guides were developed in English, translated in Luganda and tested. We recorded natural behaviors of healers at their shrines, and also verified some information gathered in the interviews. We recorded all events involving engagement of spirits and at-times, observations extended through nights for spirits ritualistic activities.

The key question was “How did you become a traditional spiritual healer (spiritualist)?” Responses to the iterative dialogue and meaning making of individual experiences stimulated interest, furthered investigations and encouraged shared learning, discovery and compared understanding without judgement (62,63). Probing questions were shaped by emergent issues during interview, in flexible unrestricted style to elicit subjective world-views of lived personal experience, constructed interpretations of their social reality and contextual meaning (64). We had flexibility to restructure questions, and managed to get personal healer information, overcame any resistance and all questions were responded to. The process clarified many misunderstandings and improved confidence of study participants. Conversations were audio recorded and conducted in local dialect, Luganda, the local language of the research participants. Recorded audios were transcribed in English while maintaining depth of context in Luganda. All transcripts were reviewed and independently coded by three researchers, whose coding were compared, merged and agreed upon by the whole research team.

### Data Management and Analysis

Collected and transcribed data was duplicated and progressively analyzed using ATLAS.ti 8 and later upgraded to ATLAS ti. 22 computer software. An initial analysis of responses was done after first three sets of semi-structured interviews. Every subsequent interview was compared with previous analysis for similarities and differences. Data was merged and distinct inconsistence and disagreements redone and cleared by the research team.

### Ethics consideration

Ethical approval of this study was sought and obtained from Research and Ethics Committee at Mbarara University of Science and Technology (No. MUREC 1/7) and Uganda National Council of Science and Technology (SS 4947). The study was also cleared by Traditional Healers’ Association (NACO/0485/2019), and Buganda Kingdom. Written prior informed consent was obtained from individual subjects.

## Findings

### Socio-demographic characteristics of respondents

Traditional healers are divided into three subcategories; traditional herbalists knowledgeable in use of plants without being possessed by spirits *(abanozi b’eddagala),* traditional spiritual healers guided by natural spirits and natural forces majorly anchored in mountains *(Balutansozi),* and traditional spiritual healers guided and get possessed by ancestral spirits (*Balubaale)*. We were interested in spiritual healers engaging ancestral spirits, as spirit medium, during health management. Therefore, the study focused on *Balubaale*.

Study participants were aged between 27-77 years (Mean age 54), from 8 counties, 11 Districts, and 9 clans. Two doubled as *Balubaale* and *Baluutansoz*i and eight belonged to traditional religion. Two never attended school, 6 never completed primary level and 4 completed secondary level education. All respondents belonged to healers’ associations, had more than 10 years of experience and were subsistence farmers.

Age, gender, background and environmental factors contribute to individual process of becoming spiritual healers (65,66). Ratio of male to female was (5:1). Traditional spiritual healing is associated with variable gender specific restrictions, enshrined in socially constructed community taboos. Men, historically and culturally dominate traditional spiritual healing services, and women are comparably greater spiritual healers in reproductive and domestic issues (67). Female medium with dominant male spirits are not normally married because they are spiritually married to spirits (67). In this study Spirit *Muwanga*, part of criteria for inclusion of respondents, is a male spirit and that could partly explain the fewer female participants. This article focused on the detailed process of becoming a spiritual healer (*Malubaale)*. Participants were asked an open-ended question “*How did you become a traditional spiritual healer/spiritualist (Mulubaale)?”*

In response, participants described becoming a *Mulubaale (Singular)* as an involuntary sequential five-to-ten years process that involve participation of family, relatives and clan members. To many, the involuntary process started as an unknown gift manifesting as multifaceted problems involving illness in an individual or within family that resolved through cultural and spiritual rituals guided by ancestral spirits. For example, one participant explained “… *it was not my wish to become a spiritualist. I was forced into spirituality because of illness. I turned out to be mentally ill and become mad”*. (Participant 4 [M, 60’s]). Participants disclosed that to become a *Mulubaale*, an individual goes through four major steps. First step is identification of signs and indicators of anointment. Second step is exploration of ancestral spirits (*kwaza Lubaale*) within the family. Third step is training and apprenticeship (*kutendekebwa*) of spirit medium (*Mutende*) and their ancestral spirits (*Lubaale)*. Fourth step is pass-out or graduation ceremony attended by community members.

### Identification of signs and indicators of anointment

Spiritual healing is an inherited ability that may manifest as illness that fails to respond to conventional treatment and/or with indicators of a natural gift that require to be understood and nurtured. Some participants reported being born gifted and imbued with healing abilities that flow smoothly and naturally for them. A participant narrated *“I was born the day my great grandfather (name removed) died, yet my mother had stayed pregnant for three years. So, I was named after my great grandfather and I also inherited his healing abilities”* (Participant 1 [M, 40’s]).

Participants stressed that anybody cannot become a *Mulubaale,* unless he/she has been identified and called by ancestral spirits. They have indicators such as birth marks, recognizable by elders or experienced healers during infancy. One healer indicated that by age of five years, he was already offering healing services to family and community members. He would get possessed by ancestral spirits, assess illness and prescribe remedies. One participant commented “*Ancestral spirits select and prepare their spirit mediums.”* (Participant 12 [M, 20’s]).

Some participants reported being gifted in accessing health care information through clear dreams of clients, descriptions of plants to use, and clear instructions on preparation and dispensation of medications to clients. Two participants reported heightened sensitivities to light, sound and vision. They access information through hearing active voices, seeing images of people giving detailed instructions on medicines to prepare. However, others claimed to “just know”. They reach a level when they know without dreaming or getting possessed by ancestral spirits.

People unaware of their gifts or fail to nurture them suffer with physical manifestations of illness. as individuals or their family members. madness (*eddalu*) For example, one participant reported “*… I started experiencing indictors of becoming a spiritual healer at childhood years. The indicators included illness, dreams and seeing images of people or animals (obutulume) which other people cannot see.”* (Participant 2 [M, 70’s]). He got healed when the explored their ancestral spirits.

### Exploration of ancestral spirits (*Kwaaza Lubaale*)

*Kwaaza Lubaale* is a cultural ritualistic process of exploring ancestral spirits, is often prompted by illness and engages many family members. It serves to invite and identify ancestral spirits and individual human spirit mediums among family and clan members. One participant explained. *“… multiple problems and illness in the clan forced us to explore our ancestral spirits. We were more than 80 people, and amongst all the people, ancestral spirits possessed me”* (Participant 9 [M, 50’s]). Exploration of ancestral spirits is divided into three major steps undertaken in a particular order. First, preparatory steps. Second, dressing ancestral spirits that have expressed the need to be dressed (*okusumikira empewo*). Third, throat cutting of animals identified by ancestral spirits for sacrifices (*kusalira)*.

### Preparatory steps preceding exploration of ancestral spirits

Exploration is preceded by three major preparatory activities intended to normalize ancestral spirits. First, to visit, clean and pay tribute to ancestral spirits in family grave yards (*okulima ebijja*). Second, rituals to harmonize clan and family twins and twin forces *(okwalula abalongo*). Third, to perform pending last funeral right for dead clan/family members (*okwabya enyimbe*). For example, one spiritualist said *“… we prepared ourselves before the ancestral exploration, properly cleaned the family grave yards, performed the pending last funeral right for the dead, and harmonized rituals for the twin sets”* (Participant 5 [M, 60’s]). At the end, family members gather and undergo cleansing rituals (*kwambulura)* to prepare them for inceptive stage.

### Inceptive stage of *kwaaza lubaale*

An adult male black goat and a multi-colored male chicken, resembling an owl (*enkoko ya kiwugulu)* are put in place after the cleansing ritual, and a to attract and harmonize ancestral spirits as stated by one participant. a communal meal (*ekijjulo*), involving various types of local foods, fruits, animal products and local drinks is prepared and served amidst singing, drumming and dancing. The essence is to persuasively attract various ancestral spirits to express themselves and possess their preferred family members as was expressed by one participant *“… as the family gathered, a black male goat and an owl-looking male chicken are put in place to attract the spirits together and remove any conflicts therein.”* (Participant 11 [M, 50’s]).

The ancestral spirits that express themselves are welcomed, carefully interrogated and evaluated, one by one, by lead spiritualist (*Ssenkulu*) and elders, in presence of family members to articulate other details such as, spirit’s name, its family lineage of origin, artefacts, regalia, its lead *Muzimu* who owns the ancestral spirits and the name of the person who last harmonized these spirits. Drumming and singing would continue for three consecutive sets of nine days or until *Muzimu,* appears, which closely associates with the ancestral spirits it owns and controls. When other ancestral spirits, *Misambwa (*superior mystical ancestral spirits), and *Mayembe (*worker/assistant ancestral spirits) appear, each is probed for identity, characteristics, functional roles and their main *Muzimu*. For example, one participant said; *“… during spirits exploration, every spirit that appears is probed for its details regarding its origin, roles, responsibilities, regalia and its lead Muzimu”.* (Respondent 6 [F, 60’s])

At this moment, we can give a summary of our observation. Most spiritual gatherings started in evenings, took place indoors and involve many family members. Lead spiritualist (*Ssenkulu*) was always assisted by his trained relatives or solicited assistants from other invited spiritualists. Communal drumming and singing were associated with recitation of guiding myths, riddles and instructive stories that lasted for several hours. *Ssenkulu* diligently observed the family members who become possessed and provided necessary space for what would follow. An individual would suddenly start dancing intensively, then get possessed, stroll and roll on the ground while shrieking, panting, and twisting the head in many directions. *Ssenkulu* would carefully communicate with the ancestral spirits. Some spirits that started by using threatening abusive language, would gradually be cooled down by the sober *Ssenkulu* who performed many rituals in the process harmonizing with the spirits. Finally, the evaluated ancestral spirits demanded that *Ssenkulu* dresses their human spirit medium.

### Dressing the human spirit medium (*kusumikira*)

*Ssenkulu* has the responsibility to dress the human spirit medium when the family members and himself are satisfied with the expressions of the ancestral spirits of dead persons (*Mizimu),* superior mystical ancestral spirits (*Misambwa)* and worker/assistant ancestral spirits *(Mayembe)*. For example, we witnessed a male teenager human spirit medium being dressed with three knotted backcloths. *Ssenkulu,* dressed in his regalia and got seated on animal skin (*ekiwu*), facing in-front of the boy who was seated on a new sheet of backcloth (*omwaliiro*), in presence of the boy’s father. A male assistant spiritualist placed the already gathered materials between the *Ssenkulu* and the boy. The materials included three knotted backcloths, two baskets (*bibbo*), a walking stick, a spear, and many plants species. *Ssenkulu* suddenly got possessed by a male spirit which introduced itself as *Kawumpuli* and narrated its royal ancestral lineage including the details of its father, King *Kayembe,* its mother *Nakku,* its guardian mother *Nabuzaana* and the details of miserable childhood, upbringing, and how it became a Prime Minister (*Katikiro*) of all ancestral spirits of Baganda. (We were advised to exclude greater details). The *Kawumpuli* spirit introduced its medium by name (withdrawn), his paternal and maternal ancestral clan lineages and their burial grounds. *Kawumpuli* detailed the basis of that shrine (withdrawn on request), and made an assertive statement, *“You (Muzimu’s name), in case you are not comfortable with me (Kawumpuli spirit) and this shrine, feel free to express yourself in any way and we shall stop immediately.”* (Spirit of participant 6 [F, 60’s])

Instantly, the boy got possess by a *Muzimu* spirit that mentioned its name (withdrawn) and confirmed that the process should continue. The spirit left the boy soon after its affirmation. The *Kawumpuli* spirit instructed its human male assistant to introduce his name, the details of his ancestral lineage, including the names of his parents and their respective clans. Our request to audio and video record was granted by the spirit. *Ssenkulu* stood up and dressed the boy spirit medium with three knotted bark-cloths. The first knotted backcloth for *Muzimu* was placed facing on the right side of shoulders of the boy. The second knotted backcloth for *Misambwa* was placed facing the left side of the shoulders, and the for *Mayembe* faced the right side on top of the first one for *Muzimu*. The process that follows is throat-cutting (*Kusalira*).

### Throat-cutting for ancestral spirits (*Kusalira Lubaale*)

Throat-cutting for ancestral spirits is a cultural ritual done for ancestral spirits *Mukasa*, *Kiwanuka* and *Musoke*. Goats, sheep and chicken of specified sex, maturity and colors are slaughtered in a process spearheaded by ancestral spirit *Muwanga* using his particular knife (*ekyambe kya Muwanga*). The rituals for *Lubaale Mukasa* and *Kiwanuka* are done on the same day at about 5.00 am in the morning. The rituals for *Lubaale* Musoke are done the next day at about the same time, 5.00 am. The throat-cutting rituals for *Mayembe* are done periodically, as agreed upon between the spirits, spirit medium and family members. The throat-cutting process serves to strengthen the spiritual powers of ancestral spirits. One participant narrated *”… throat-cutting is done by spirit Muwanga to empower and rejuvenate ancestral spirits Misambwa and Mayembe”* (Participant 10 [M, 40’s]). Finally, the identified ancestral spirits are introduced to the clan or family shrine and the *Ssenkulu* to lead their training is agreed upon.

### Training and apprenticeship of spirit medium (*kutendekebwa*)

The identified human spirit medium undergoes structured 5-10 years mandatory training and apprenticeship process that takes place at the *Ssenkulu’s* shrine. The ancestral spirits are called in one-by-one and categorized into *Mizimu, Mayemb*e, and *Misambwa*. *Misambwa* are further sub-categorized into dryland spirits that include *Muwanga*, water-related spirits (*Balunyanja*), and royal-spirits (*Balangira)*. During training, the characteristics and health management roles of junior ancestral spirits are stated, harmonized and explained senior ancestral spirits. For example, one participant said, *“…superior ancestral spirits are responsible for preparing the newly expressed ancestral spirits,* (Participant 7 [M, 50’s]).

The ritualistic training is intended to prepare, guide, purify, and empower ancestral spirits. Trainees are closely supervised while using ancestral spirits to assess, diagnose and manage illness of clients at *Ssenkulu’s* shrine, until supervision gradually reduces to minimum. Knowledge and skills acquired are influenced by ancestral spirits of the trainer and the trainee. One participant said *“… I learnt from my training that we train same basic principles of using ancestral spirits for health care, but the knowledge and experience varies depending upon the trainers, their ancestral spirits and the ancestral spirits of the trainee.”* (Participant 3 [F, 60’s]).

Training does not follow particular chronological order, but serve transition of ancestral spirits into being functional, establishes connection with spiritual powers of living and non-living materials, non-materials, and sacred places through rituals. Trainees learn about dreams and dream-interpretations, relate symbols and symbolism and their application in health care, understand the significant of regalia for various ancestral spirits, composition and use of diagnostic tools (*mweso*), and the essence of ethics and professional code of conduct.

### Transitioning ancestral spirits into becoming functional

Transitioning newly expressed ancestral spirits into function is through rituals performed on the spirit medium and spirits undergoing training. We observed trainees fell down as they get possessed by ancestral spirits in the beginning, termed *“Lubaale omuto agwa.”*. After repeated counsel and rituals by *Ssenkulu* the ancestral spirits anchor properly on the host’s head without falling down, *“Lubaale omukulu tagwa”.* Harmonized ancestral spirits respond appropriately to simple conversations with *Ssenkulu.* Participants articulated the need to train and harmonize both the ancestral spirits and spirit medium, since they belong to old and new generations respectively. Both have to learn from each other’s reality since they have variable experiences, values, environments and structural set ups which need to be synchronized. *“Whenever ancestral spirits possess a new human spirit medium, they are in a new human generation and environment both of which need to know and learn from each other.”* (Participant, 12 [M, 20’s]; and 2 [M, 70’s])

### Connecting with spiritual powers of material and non-materials

Connecting with spiritual powers and energies was subcategorized into spiritual powers of living and non-living materials, non-materials, and sacred places. Trainees are initiated and taught how to connect with and use spiritual powers of living things such as plants, animals, birds, reptiles and insects (details removed on request of study participants). They also to use spiritual powers of non-living things such as waters of lakes, rivers, springs and rains, rocks, and parts of dead plants and animals. One participant concluded *“I came to understand how to use medicinal materials after the initiation and during my training and apprenticeship.”* (Participant 11 [M, 50’s])

In addition, trainees learn to connect and use the complex cosmology, tap into energies of the sun, moon and stars. Space-time relationships of the universe are highlighted. One participant said *“I get surrounded by and connected to the powers of the Sun, Moon and Stars, and I am able to utilize them for healing.”* (Participant 4 [M, 60’s]). Furthermore, participants explained that they are excluded and taken in isolation where they explore spiritual powers of nature and sacred places such as mountains and forests. They learn to use spiritual powers of ritual fire for healing. (Details excluded). Various rituals are performed for trainees to experience the spiritual powers generated through individual and communal rituals and the significance thereof.

Communication with ancestral spirits is very important for spiritual healers. Ancestral spirits communicate through dreams, intuition, spirit possession, illusionary sounds (voices) and images (visions). Communication is facilitated by use of music and drumming, local brew, fire (*Ekyoto),* incense (*kabaane*), roasting meat, pipe smoking, or token of money *(kigali*). For example, ancestral spirits who used to smoke a pipe may demand their medium to smoke pipes of particular nature, shape, color and content. Specific rituals are done on particular pipes to ease communication. Trainees are further taught how to communicate to categorized ancestral spirits through fire and fire places, singing and praying with contextualized information.

### Dreams and dream interpretation

Skills in dreams and dream interpretation are significant for spiritual healers. Trainees share dreams amongst themselves and with *Ssenkulu, and* are encouraged to understand, interpret and analyze dreams and the constituted ancestral communications. Dreams are consciously related to symbols, colors, living and non-living things, and associated ancestral spirits for connectivity and meaning. For example, one participant illustrated *“I used to dream of an old man requesting me for his shoes. I bought many pairs of ordinally shoes from our nearby market, but the dream was persistent until I shared with my Ssenkulu. Ssenkulu interpreted that my grandfather was ready to use his diagnostic tool (omweso gw’engatto), and dream stopped when diagnostic tool was constituted, the dream ceased,”* (Participant 1 [M, 40’s])

### Utilization of symbols and symbolism

During apprenticeship, trainees are guided through understanding symbols and related symbolism to various illnesses, colors and associated ancestral spirits which are utilized during health management. For example, one participant explained *“… at childhood age, I was chronically ill. My illness was interpreted and related to ancestral spirits. The illness was explained to mean that I was to become a spiritualist and I would never recover from the illness until I start on my spiritual journey of becoming a traditional spiritual healer.”* (Participant 2 [M, 70’s])

### Training in regalia and its significance

During apprenticeship, trainees acquire competence in regalia characteristics and significance for various ancestral spirits. *Ssenkulu* discusses the restrictions, taboos and rituals required to maintain regalia sacred. Regalia refers to distinctive traditional ritualistic clothing and ornament items of cultural vitality that symbolize rooted relationship and communication between the human, spiritual and natural worlds. We observed regalia of various types, shapes and colors placed in special places within participants’ shrines. There were several baskets (*bibbo*), reeds, spears (with one or multiple heads), symbolic knife for Spirit *Muwanga (Empiima/ Mwambe*), traditional rosary made of large black beads *(Lutembe),* authority stick *(Ddamula),* seat-mat made out of skins of sacrificed animals (“*kiwu”* for singular*; “biwu”* for plural), smoking pipes (*mindi*), drums, spiritual harmers *(enyondo),* knotted bark-cloth *(Ekifundikwa),* walking stick (*omuggo*), among many others.

### Training to use diagnostic tool (*mweso*) and its composition

Trainees learn divination and to use the diagnostic tool (*omweso*). *Ssenkulu* performs rituals that facilitate communication between the trainee, the ancestral spirits and the diagnostic tool. The diagnostic tool is constituted and empowered by *Ssenkulu* under the guidance of ancestral spirits of the trainee owning the diagnostic tool. When the diagnostic tool is fully constituted, and the trainee has learned the basics of how to use it, it is then tested and approved by the ancestral spirits possessing the trainee by rightfully divining the *Ssenkulu*. For example, one participant narrated *“… the muzimu of my grandfather possessed me and requested Ssenkulu to constitute its diagnostic tool. My ancestral spirits guided the Ssenkulu in the process of constituting and empowering its diagnostic tool. At the end of the process, the Ssenkulu handed over the constituted diagnostic tool (Omweso), to my ancestral spirit (Muzimu) and asked it to divine him, if my it was satisfied with the work so far done. When the Muzimu spirit rightly divined the Ssenkulu, Ssenkulu was satisfied with the divination, because the divination was real.”* (Participant 6 [F, 60’s]). It was noted that the divining tool is best used by its owner and his ancestral spirits.

### Training in ethics and professional conduct of spiritual healer (*Mulubaale*)

Ethics and professional conduct are an unwritten integral part of the entire training and apprenticeship process. The dos and don’ts of the career are highlighted, and the aspects of sexual discipline are stressed. Times to abstain from sexual relationships are specified and emphasised. Trainees and ancestral spirits are guided how to avoid being ill-advised into wrong doing, and the expected repercussions upon violation of ethical conduct are elaborated.

*Ssenkulu* is ethical, competent, experienced and knowledgeable, and is highly respected by ancestral spirits. He has authority and capacity to discipline ancestral spirits, address their undesired acts and mediate any conflict between the ancestral spirits and their human spirit medium. One participant stated *“… my Ssenkulu disciplines my ancestral spirits, and mediates conflict between me and my ancestral spirits.”* (Participant 8 [M, 30’s]). Training and apprenticeship follow strict cultural norms and ancestral roots, and mistakes are avoided because it is very expensive to rectify a mistake in spirituality. The ancestral spirits are advised to be prompt when called upon, cautioned not to engage in humanly issues relating to their human host, and never to involve themselves in ceremonies that did not concern them (as spirits).

### Training in Health Management

Trainees are guided to become skilled and competent in health management. They are trained to understand signs and symptoms, to do health assessment, make spiritual diagnosis, treatment, and related preventions. They are expressly taken through causes of illness by various ancestral spirits, cultural issues and witchcraft. They engage ancestral spirits and cultural materials in an orderly manner during health management. One spirit is allowed to engage the human spirit medium at a time. Particular roles and responsibilities of various ancestral spirits are clarified. When *Ssenkulu* is satisfied with the trainee, the ancestral spirits, and that they can work independently with minimum supervision, he informs the trainee and their relatives to prepare for the pass out and graduation ceremony.

### The pass-out and graduating ceremony

The last aspect of training and apprenticeship spiritual healer is the pass-out and graduation ceremony which has three noteworthy aspects. That is, the final assessment, the communal rituals and community witness.

### The final spiritual assessment

The final assessment trail is locally referred as ‘*akasera ka Lubaale’.* It is the last spirituality examination winding up the training and apprenticeship of the spiritual healer (*Mulubaale*). It is done in presence of known experts and experienced categorized spiritualists, who give independent marks and consented grade to trainees. For example, one participant said *“I did my final examination at Buddo in presence of four prominent and specialized spiritualists (names withdrawn).”* (Participant 4 [M, 60’s]). The process is carried out at night in presence of selected people. The research team was privileged to attend, but was instructed not to disclose the details. Those who fail the final assessment are advised to master the skills and are advised to be considered another time.

### Pass-out and graduation Communal ritual

Pass-out and graduation is a communal celebration associated with communal meals that are contributed to by family, clan and community members. Animals and birds are slaughtered in addition to those instructed by the ancestral spirits. Family and community members are fully involved in the whole process. Their involvement bears witness to the authenticity of the graduating trainee. It also serves to facilitate acceptance of the trainee as a spiritual healer in the community. One participant said that the presence and participation of the community members is very important, since no certificate is given out to mark the successful completion of the training and apprenticeship. The participant narrated *“I was very excited to see all my family and community members attending my pass-out and graduation. I felt accepted as a competent spiritual healer (Mulubaale). However, I was disappointed that I was not given any certificate.”* (Participant 3 [F, 60’s]).

### Community witness

Pass-out and graduation is very important community activity witnessed by many people including experienced spiritual healers, other trainees, family members, local and religious leaders, members of the clan and the entire community. They all bear witness to the successful completion of the spiritual healer trainees within their respective communities. One participant said “…*over 250 people attended my graduation. Family and community members mobilized everything including cows, sheep, goats, chicken, local brew and backcloths for the ceremony.”* (Participant 11 [M, 50’s])

### Genuine and authentic Baganda spiritual healers

Participants stressed that genuine and authentic Baganda traditional spiritual healers (*Balubaale abatukiridde*) are characterized by their naturally embedded gifts and trainings. They are ethical human spirit mediums whose graduation was witnessed by their family and community members. They are connected with their ancestral spirits and sacred places, and are trusted and confided in by their family and community members. Their regalia and divining tools are reflective of their major working ancestral spirits in line with their natural gifts and training. Their healing abilities are based more on their healing hand *“omukono oguwanya”* than the medicine they give and whatever they undertake in healing flows smoothly.

Participants listed three coordinated characteristics to identify genuine and authentic Baganda traditional spiritual healers (*Balubaale*). Firstly, the dressing regalia of the human spirit medium and content of their immediate working environment. For example, *“The spirit medium for Kiwanuka is dressed in red”.* (Participant 9 [M, 50’s]). Secondly, they are acknowledged as community resource members by the community where they work and reside. One participant narrated *“… I am proud of this community and the community has been proud of me and my works for the last 20 years. Most community members are my friends and clients, and most youth are my grand-children. I have helped families to stabilize, conceive, assisted with antenatal and postnatal care and I have conducted most of their deliveries. … I am consulted for guidance in everything.”* (Participant 5 [M, 60’s]). Thirdly, they are motivated by the success of their practices. One participant explained *“It is very motivating for me to successfully manage or treat very difficult health conditions. I find interest in handling cases which others have failed. I am interested by clients who trust me and my work.”* (Participant 11 [M, 50’s])

## Discussion

Using an open-ended question “how did you become a spiritual healer?” the findings show a five-to-ten years guided process that often starts in response to an illness. Spirit mediums are identified to undergo mandatory structured training where their ancestral spirits are explored, categorized, and ritualized. Both, the human spirit medium and the ancestral spirits learn how to connect and communicate with each other and with the spiritual powers of other living and non-living materials, non-materials, sacred places and the spiritual powers generated through rituals. They also train in dream interpretations, symbols and symbolism, regalia and their respective significance, traditional diagnosis, and ethical conduct during health management. Successful completion of training concludes with final assessment (*akasera ka Lubaale*), communal rituals and a graduation involving stakeholders, family members and witnessed by the community.

Previous researchers documented similar findings, that spiritual calling into healing is an intergenerational familial natural gift (16,23,68). Another researcher found that anointment into healing using ancestral spirits is involuntary, and may manifest through challenges of illness (69). For example, the incurable mental disorder that only responds to treatment after the individual is initiated into healing was expressed below.

> *“What I know is that some of the mental health problems in children and young people is caused by ancestral clan spirits, especially if these spirits want the person initiated into being a traditional healer and the person resists… and not until this person is initiated into traditional healing his mental disorder never heals” – Traditional healer 13, (53)*

Therefore, illness is not always acknowledged as harmful, since some people may be stricken by particular illness, associated with ancestral spirits, to become healers (70).

It is also documented that initial preparatory rituals facilitate ancestral spirits communication through dreams and visions (71), throughout spiritualists’ training and apprenticeship (72,73). Therefore, gifted spiritual healers are privileged to access mythical and sacred realms of community traditions, imagery powers, reincarnation, spirits guide and participate in experiences transcending human life.

According to Abbo, the methods of departure from Western medicine are underpinned by training and practices (74). Unlike Western formal models of medical training where emphasis is put towards the power of using the five sense organs of tough, smell, hearing, testing and seeing, the learning of spiritual healers is imparted to the young generations by elders as an integral part of community’s social, spiritual, ancestral and natural environment (75). The training engages the presence of specialized spiritualists who encourage the trainees to be aware and conscious of, and experience situations beyond the power of sensing or using one’s intellect, into a spiritual zone where one just observes and witnesses what the mind does, without thinking. This repetitive training is a practice-knowledge interface that synergizes the strength of practice and knowledge (76), towards self-transcendence where self is connected to the collective spiritual, emotional, cultural and psychological being as well as a physical with material embodiment (77). Other distinct aspects of the spiritual healers’ training are that apprenticeship take place within the community, their training is verified by people who are communally recognized and acknowledged to have the necessary expert knowledge and skills (72), and their graduation is witnessed by cultural, religious and local leaders, family and community members, which partly account for the high trust and confidence people have in them (69,72).

Although the ways spiritual healers obtain their knowledge and skills vary widely (23), and the reliability of ancestral communication and its contextual packaging is highly arguable, the valid sources of exact knowledge stand questionable (78). Some countries, such as China, Korea, India and Vietnam, have established official education/training programs for traditional medicine practitioners at university levels (13,18). In Uganda medical students support inclusion of the traditional medicine principles in their medical school curricula to understand the traditional healers paradigm and foster collaboration (79). Even then, spiritual healers are still not recognised. However, some authors have reasoned that personality and appropriate use of words and rituals create necessary vital forces required for therapeutic impact (44).

Traditional healers mobilized themselves into associations to stay afloat amidst unfavorable regulatory frameworks (80). In Uganda, PROMETRA Uganda and THETA NGOs worked together with government agencies towards actualization of the Traditional and Complimentary Medicine Act 2019 that recognizes health care activities of traditional healers in Uganda (43). Academic and research institutions such as Makerere University, Mbarara University of Science and Technology (PHARMBIOTRAC), Gulu University, Dr. Sekagya Institute of Traditional Medicine (Dr-SITM) (https://dr-sitm.com/), National Agriculture and Research Organization (NARO) and National Forestry Authority (NFA) have strengthened positive engagement with traditional medicine and healers (23,79).

### Study limitations

This study was limited to one category of traditional spiritual healers (*Balubaale*) who use ancestral spirits during health management, which limited the scope of data from other categories of traditional healers. Limiting the study to Central Uganda may have limited the generation of data and more insightful conclusions. Although these limitations affect the generalizability of the findings, the findings may not differ much in context for other tribes elsewhere in Uganda.

### Conclusion and recommendation

This qualitative study is the first of its kind to document the detailed process of becoming a traditional spiritual healer using ancestral spirits in health management (*Mulubaale*) among Baganda. Becoming a *Mulubaale* is considered a special gift for service, where an anointed person explores and connects with their ancestral spirits and undergoes supervised training and apprenticeship by knowledgeable and experienced senior spiritual healers. Their apprenticeship and graduation are witnessed by cultural, spiritual, religious and local leaders, family and community members. It is worth mentioning that these spiritual healers are important within the health system as a bridge between formal institutional care and their cultural context. This article contributes to existing literature on training of traditional spiritual healers and forms a basis for contribution, discussion and further research regarding formalizing traditional spiritual healers as health workers in Uganda.

We recommend similar studies to be conducted within other ethnic groups of African spiritual healers to understand the similarities and differences of their practices. This will facilitate common understanding and harmonization around traditional spiritual healers’ practice and contribute towards health care. We also recommend to have intercultural health approach in institutional care like it is in Chile and Peru, to concretize these findings and contribute towards appropriate policy considerations.

## Data Availability

All data produced in the present work are contained in the manuscript

## Acknowledgement

We extend our gratitude towards the leadership of Mbarara University and the PHARMBIOTRAC program for the financial support towards the PhD study of the first author. Particular thanks to Dr. Casim Tolo, Professor Patrick Ogwang and Engineer Anke Weisheit. We thank the study participants and their ancestral spirits for their committed participation in the study and for the information shared. We are equally grateful to the leaders of traditional healers’ associations, namely Sylivia Namutebi (Maama Fiina) of Uganda N’eddagala Ly’ayo N’obuwangwa Bwaffe, Musasizi Karim of NACOTHA, and Karim Walyabira of Uganda N’eddagala Ly’ayo. We acknowledge Oweky. Kyewalabye Male of Buganda Kingdom for introducing us to the clan leaders. We thank the Baganda clan leaders and district leadership for your support in accessing the study participants. We thank the research assistants Mr. Eric Kibirige Mukasa and Ms. Nambuya Barbra for the field work. We also thank Dr. John Chrysostom Katongole, Jovent K. Obbo of Bugema University and Dr. Kizito Simon of Makerere University for their support.

## Supporting information

All the data supporting our findings are contained in the paper.

## Contextual Definitions (Legends)

In the context of this study, the definitions or legends below are based mainly on the field findings.

**Ancestors** refer to persons that have passed on to other dimensions of existence, for whom people are descendants. They include relatives such deceased parents, grandparents, great-grandparents, aunts and uncles, animals, birds and stones

**Ancestral spirits** refer to guardian spirits that possess people and talk through them as medium. They also guide through dreams and these include *Mizimu*, *Mayembe* and *Misambwa*.

**Animals** in the context of this study, refer to low-conscious living things in form of animals, reptiles, insects, birds, etc. empowered with spiritual powers which are used in health care by the traditional spiritual healers

**Apprentice** is formal hands-on-training, a traditional spiritual healer trainee undergoes, while staying and working closely with a senior specialized spiritualist, to learn how to interact with the ancestral spirits, to know and use ancestral spirits to manage and heal cultural and spiritual health issues.

**Baganda** (singular *Muganda*) are an ethnic group of Bantu speaking people of Buganda kingdom in Central Uganda, with a recognised hereditary leader known as the Kabaka. They consist of clans, sub-clans and families linked by social and blood ties, with a common culture, and speak *Luganda*. as their language.

**Beliefs** refer to what Baganda traditional spiritual healers thought and trusted as true or correct with or without guiding reason to one’s decision, action or behavior.

**Diagnostic tools** refer to a set of objects, upon which rituals are performed, used by traditional spiritual healers to understand the nature of problem, its root cause and the ways to manage the problem. For example, the use of coffee beans and cowrie shells.

**Disease** refers to affected body parts that manifest signs, symptoms such as pain and fever. **Divination** refers to cultural process to foretell and give advice based on retrospective and prospective foresight by traditional spiritual healers

**Dreams** are images, ideas, feelings or thoughts that flow while one soul travels to the spiritual realms when the body is sleeping.

**Health** refers to the resultant situation of the harmonized and integrated human life with other low-conscious living things, the environment, and the sociocultural, spiritual and metaphysical factors in nature.

**Health care** refers to socially acceptable efforts made to maintain or restore the physical, mental, emotional, psychological, financial, spiritual and social well-being of individuals, families or community.

**Illness** refers to the subjective ways practitioners understand the manifestations of the affected soul and spirit parts, the root cause, and management.

**Initiation** is a rite of passage spiritualists go through that involve spirit-specified rituals and tests to confirm his/her connections with the ancestral spirits.

**Intuition** is the ability for one to understand through spontaneous instinctive feelings of smell, touch, hearing, knowing, seeing without necessarily using facts, evidence nor conscious reasoning.

***Lubaale*** refers to individual or collective ancestral spirits.

***Mayembe*** (*Jjembe* singular) refer to ancestral spiritual entities headed by *Jjembe* Lubowa. They assist on various roles and responsibilities utilized as worker spirits

**Mediums** refers to animals or human beings utilized by ancestral spirits and act as intermediary channels of communication between ancestral spirits and human beings.

***Misambwa* (***Musambwa-singular*) are most superior categorized ancestral spirits with persistent impressions in the communities.

***Muzimu (****Mizimu-Plural)* is the collective consciousness of that person who passed on to the spiritual realm mainly characterized by the soul.

**Offering** refers to anything intentionally given out for ritualistic purpose towards ancestral spirits and other supernatural forces.

**Plants** in the context of this study refer to vegetation empowered with spiritual powers used in health management by the traditional spiritual healers

**Regalia** refers to distinctive traditional ritualistic clothing and ornament items of cultural strength and significance that symbolize rooted relationship and communication.

**Rituals** are recognized cultural practices rooted in tribal spirituality.

**Ritual healing** is traditional medicine practice rooted in cultural spirituality that may involve a wide range of formalities and materials used to manage the physical, mental, emotional, financial, spiritual and psychological difficulties.

**Sacred places** are dwelling places for spiritual beings, imbued with spiritual, supernatural and mystical powers and abilities. Examples include places such as natural forests, rivers, mountains, shrines, and fire-places.

**Sacrifice** refers to intentional giving of animals and/or birds for ritualistic purpose towards ancestral spirits, supernatural forces and divine beings.

**Spirituality** refers to ancestral, supernatural, inborn, involuntary and practical reality that guides people and society based on the values, meanings and relationships to the sacred by which people live.

**Symbol** is a representation of spiritual information, connectivity and power contained in a material and associated ancestral spirits. For example, ornament items, ritual cloths, and colors

**Baganda traditional spiritual healers (*Balubaale*)** are native health practitioners, able to communicate with ancestral spirits as medium, for health management of individuals, families and communities using spiritual powers embedded in plants, animals and sacred places.

**Voices** refer to verbal sound “heard” through ears

## Notes

### Competing Interest Statement

The authors have declared no competing interest.

### Funding Statement

“The World Bank and the Government of Uganda through Pharm-Biotechnology and Traditional Medicine (PHARMBIOTRAC) (P151847), an Africa Higher Education Center of Excellence (ACEII) at the Mbarara University of Science and Technology (MUST), Uganda supported this work.” The funders did not play any role in this research nor the manuscript submitted

### Author Declarations

Research Ethics Committee of Mbarara University of Science and Technology gave ethical approval of this work

